# Workforce Cost Absorption among Community Health Promoters in Fragmented Maternal Nutrition and Social Protection Systems: A Qualitative Study Across Three Kenyan Settings

**DOI:** 10.64898/2026.08.26.26361384

**Authors:** Faith Siva, Danny Nyatuka, Retha de la Harpe

**Author notes:** Corresponding author: Faith Siva (FS). These authors contributed equally to this work.

## Abstract

Community Health Promoters (CHPs) connect households with formal health services. In maternal nutrition, they provide counselling, follow-up and referrals. However, pregnant women experiencing poverty, food insecurity, and socio-cultural issues in resource-constrained settings may be unable to act on nutritional advice. While social protection could alleviate such socioeconomic issues, maternal nutrition and social safety nets operate in institutional silos, creating gaps that systematically exclude vulnerable mothers from essential relief. This qualitative study examines how CHPs navigate these gaps across three underserved Kenyan settings. We analysed semi-structured interviews of 12 purposively selected CHPs from a broader study of 75 stakeholders, using the Braun and Clarke thematic analysis framework. CHPs described recurrent gaps between household needs and resources available through formal maternal health, nutrition, and social protection systems. CHPs stepped in; extending follow-up care, brokering information, negotiating access, and spending personal resources with inadequate formal mechanisms. They experienced emotional and relational pressure from community mistrust, cultural limitations, administrative gatekeeping, digital-system failures, heavy workloads, and performance targets tied to unreliable pay. These insights reveal that CHPs act as invisible safety nets for fragmented services, taking on burdens that official programs overlook. We describe this as workforce cost absorption. Recognising this hidden contribution is important for workforce planning and for designing integrated maternal nutrition and social protection programs.

## 1. Introduction

CHPs serve as a foundational bridge between formal health services and grassroots households. As trusted, trained community members, they are support health promotion, household engagement, referrals and community-level prevention (1). As a core pillar of the national healthcare strategy (2,3), they advocate optimal nutrition, hygiene, immunisation, and breastfeeding practices, while mobilising expectant mothers to seek timely antenatal and postnatal care within maternal and child health frameworks. Hence, CHPs are at the frontline of efforts to achieve Universal Health Coverage (UHC) by expanding health access (4,5).

However, the effectiveness of frontline health promotion depends on a households’ ability to act on clinical advice. In resource-constrained Kenyan settings, persistent poverty, food insecurity, fragile livelihoods, cultural influences, and other socioeconomic constraints prevent pregnant women from adhering to the recommended nutrition regimen (6,7). Medical advice often fails to match household realities, revealing major challenges for frontline workers. CHPs are expected to improve health outcomes, yet they lack authority and funding to solve the root causes of hunger (8). This results to operational frustration, as they face aggressive pushback or quiet disengagement from families simply cannot comply without reliable access to food.

Social protection programs can address some of these constraints by aiding vulnerable households. Cash transfers, food assistance, and related interventions have been associated with improved access to health and nutrition resources. However, maternal nutrition and social protection programs are often implemented through different administrative structures, eligibility mechanisms, and information systems (9,10). Gaps between these systems make it difficult for frontline workers to translate vulnerability identification into timely access to appropriate support (11–13). For CHPs, these gaps become practical problems. A pregnant woman may be referred to a facility but lack transport. A household may receive nutrition advice but lack resources to act on it. CHPs may know a vulnerable household but not connect it to appropriate social protection mechanisms. A digital reporting system may require information that cannot be transmitted reliably because of connectivity or device constraints. In each case, the formal system may have a defined pathway, but it may not work as intended, and a frontline worker must decide what to do.

Previous research has shown that CHPs perform functions beyond their formal job descriptions. They act as service extenders, cultural brokers, advocates, information intermediaries and implementers of policy at community level (14–16). Street-level bureaucracy perspectives similarly recognise that frontline workers exercise discretion when implementing policies under constrained resources, competing demands and incomplete information (17–19). These perspectives help explain why CHPs adapt but tell us less about who bears the cost of that adaptation. The question is matters because the adaptation is often described positively; showing how CHPs find alternative ways to reach households, build trust or facilitate referrals, demonstrating resilience, commitment or innovation. Yet the resources required for this adaptation do not necessarily come from the program but from the worker.

Extended follow up, using personal finances to maintain communication and contribution towards transport and immediate needs and using personal relationships to negotiate access to services help maintain continuity of care (20,21). However, they also transfer part of the operational burden of system gaps to the worker. We describe this process as workforce cost absorption; a process through which frontline workers absorb financial, material, operational, relational or emotional costs generated when formal service-delivery agreements inadequately meet realistic needs in practice (22–24). Within the settings studied, some costs created by fragmented services were transferred to individual frontline workers. However, workforce cost absorption should not be interpreted as suggesting that CHPs routinely finance health systems or that their adaptations prevent system failure.

This issue is particularly relevant to maternal nutrition as nutritional vulnerability is inseparable from social and material conditions in which pregnant women live. A CHP may provide accurate advice while having limited capacity to address socioeconomic challenges or social protection exclusion. This places the CHP at the intersection of disconnected systems whose failures ultimately affect the household. Other issues such as household decision-making, stigma, or cultural expectations also influence how women act on nutritional recommendations and these constraints shape the work CHPs undertake and the forms of brokerage they provide (25,26). Rather than looking at frontline work as just personal effort, the study examines how CHPs respond to systematic structures and family needs, asking: How do CHPs navigate gaps between maternal nutrition and social protection systems, and what costs do they absorb in responding to these gaps across three underserved Kenyan settings?

## 2. Materials and Methods

### 2.1 Study design and methodological orientation

The study used a qualitative multi-case study to examine how CHPs navigate maternal health and nutrition intervention across different social, geographical and institutional contexts in Kenya. (27,28). The multi-case design was selected to examine frontline experiences across three settings: Kibera informal settlement in Nairobi, Kajiado and Wajir counties. The cases were not intended as statistically comparable populations, but they provided different implementation environments to examine whether common processes in frontline work were evident across contexts.

The broader study examined the lived realities and information needs across multiple stakeholder groups involved in maternal nutrition and social protection. This manuscript presents a focused analysis of the CHP component of the broader study. Although the broader study included 75 stakeholders, this analysis is limited to 12 CHPs whose roles involved direct household-level engagement in maternal health nutrition. This is because the research question concerns the work CHPs undertake at the interface between households and formal health, nutrition and social protection systems.

### 2.2 Setting and Context

The study examined three Kenyan settings to investigate if frontline adaptation mechanisms and cost absorption occurred similarly under different contextual conditions. Kibera informal settlement is a high population density study area where food insecurity, limited household resources, and overcrowded living conditions complicate maternal nutrition (29). Social protection mechanisms are fragmented with multiple Non-Governmental Organisations operating, alongside the government with limited coordination. Wajir represents the Arid and Semi-Arid Lands (ASAL) which face chronic, climate-induced food shocks and significant cultural factors that influence feeding and food cultures (30). Distance, cultural practices, and limited infrastructure affect maternal health services. Kajiado is dominated by a nomadic pastoralist context in which mobility, geographical distance and socio-cultural practices influence engagement with maternal health and nutrition services. Traditional practices such as reliance on Traditional Birth Attendants and culturally-specific dietary norms affect maternal nutrition outcomes (31–33).

### 2.3 Participant Selection

For the larger study, researchers purposively recruited 75 stakeholders in maternal nutrition and social protection. The participants included expectant mothers, nurses, nutritionists, community health promoters, social protection officers, local administrators, policymakers and program leaders in health and social protection and ICT and Academia Experts. For this manuscript, we analysed interviews with the 12 CHPs who were purposively selected for their direct experience of household-level implementation and their position to describe how maternal nutrition recommendations, referrals, social protection mechanisms and other formal requirements were translated into practice. Although the focused analysis centres on CHP accounts, we consulted interview data from nutritionists and local administrators to contextualise descriptions of CHP roles and system-level processes. These accounts were not subject to thematic analysis but provided background understanding of the broader systems in which CHPs operate.

The research team approached participants through county health structures and community health units. CHP supervisors made initial contact and the research team then contacted eligible participants to explain the study and invite them to participate. No eligible participants declined participation, and no participants withdrew from the study. No repeat interviews were conducted. To maintain confidentiality, we replaced all identifiers with alphanumeric codes. Table 1 shows the first author coded different CHPs, which the second and third authors discussed and checked iteratively. Recruitment continued until sufficient conceptual depth was achieved, when successive interviews yielded no substantial insights relevant to the study objectives. The team assessed saturation through ongoing analysis during data collection. After the eighth interview, no new codes emerged and the research team confirmed that the subsequent four interviews did not introduce new themes relevant to the research question. The team did not triangulate CHP accounts with maternal accounts or administrative records for specific analysis.

**Table 1:** Participant Identity Coding and Sample.

| Stakeholder Group | County | Participant Code | Number Recruited |
| --- | --- | --- | --- |
| **Community Health Promoter**<br>**(Community Health Worker)** | Nairobi Kibera (KIB) | KIBCHW | 6 |
|  | Kajiado (KAJ) | KAJCHW | 2 |
|  | Wajir (WAJ) | WAJCHW | 6 |

**Table 2:**
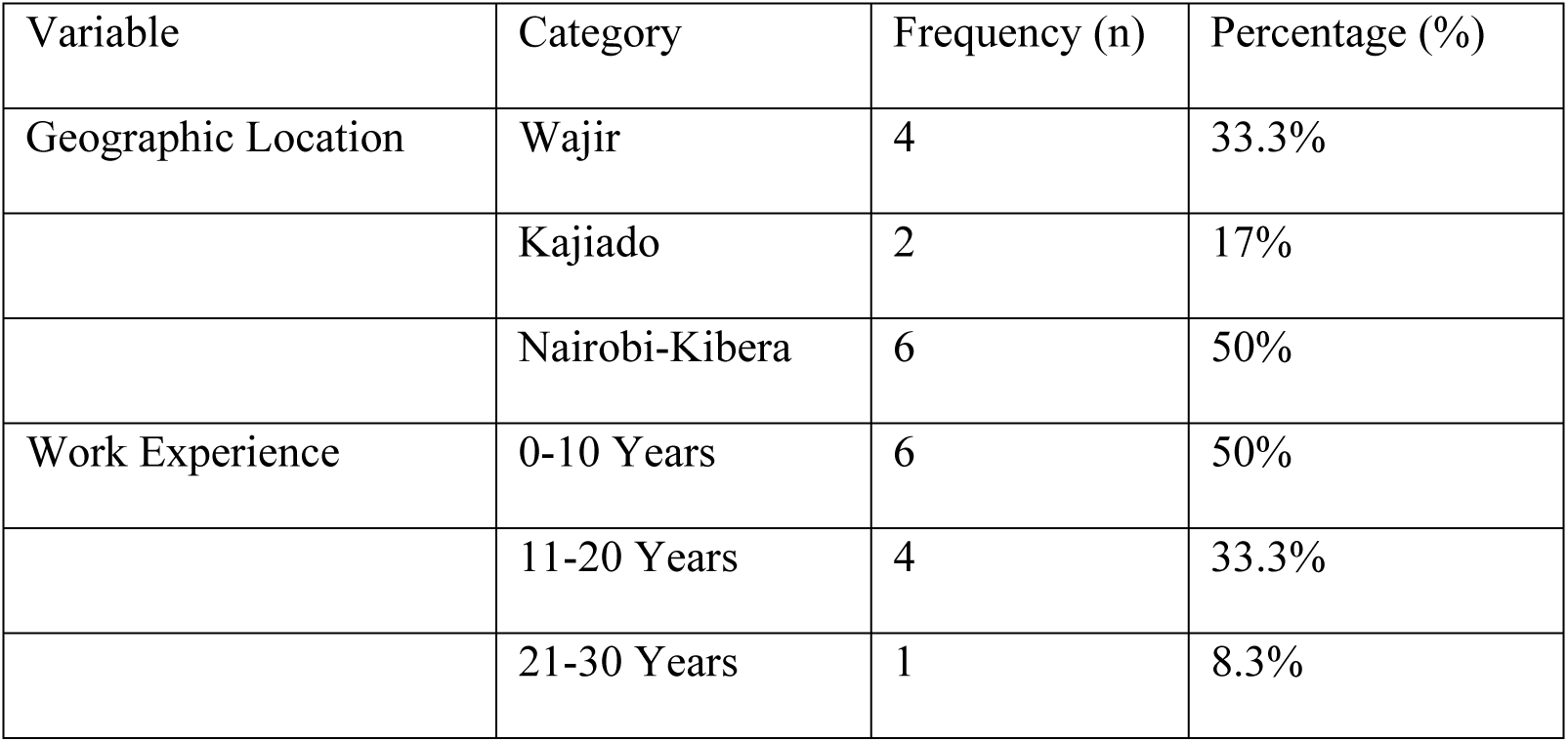
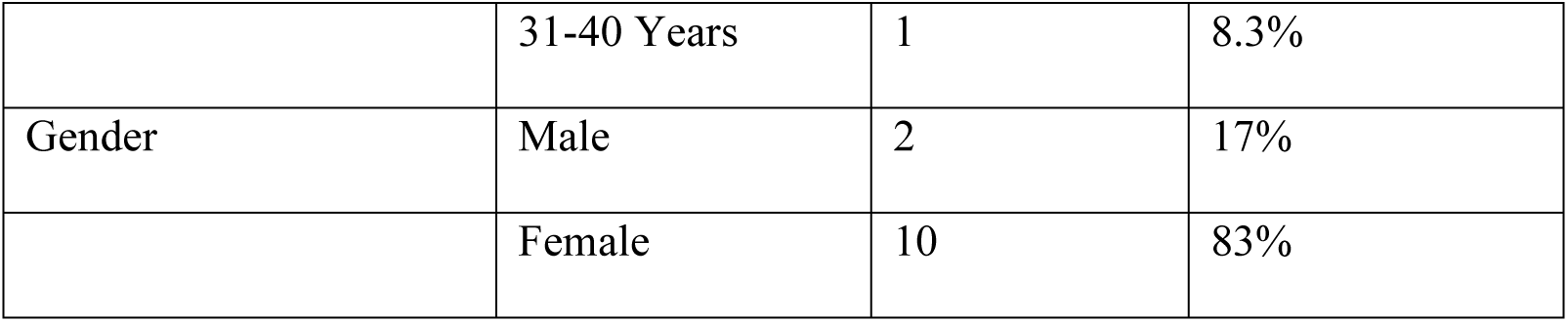
CHP Demographics (n=12)

| Variable | Category | Frequency (n) | Percentage (%) |
| --- | --- | --- | --- |
| Geographic Location | Wajir | 4 | 33.3% |
|  | Kajiado | 2 | 17% |
|  | Nairobi-Kibera | 6 | 50% |
| Work Experience | 0-10 Years | 6 | 50% |
|  | 11-20 Years | 4 | 33.3% |
|  | 21-30 Years | 1 | 8.3% |
|  | 31-40 Years | 1 | 8.3% |
| Gender | Male | 2 | 17% |
|  | Female | 10 | 83% |

### 2.4 Data Collection

Data were collected through semi-structured, in-depth interviews, in locations convenient to the participants. Interviews lasted approximately 45 to 60 minutes, and followed protocols designed to elicit detailed narratives about day-to-day operational realities. The team maintained detailed field notes were maintained concurrently, documenting contextual dynamics and behavioural observations. Some participant requested that their interviews not be recorded. The audio-recorded interviews were transcribed verbatim. The interview guide was pilot-tested before formal data collection to improve question relevance, clarity, and flow. Participants provided informed consent, and the team transcribed the interviews verbatim. The team validated interpretations by reviewing field notes with respondents to ensure accuracy. Where necessary, the team translated interview data into English before transcription. Hardcopy files were secured in physical storage while transcripts were maintained in password-protected repositories accessible only to the research team.

### 2.5 Researcher Reflexivity

The first author, a female doctoral researcher with experience in digital health, maternal nutrition, health systems, and digital transformation in Kenya conducted data collection. She has training in qualitative interview methods and has previously conducted research with CHPs in Kenyan settings. Before the interview began, participants were informed about the study objectives and the researcher’s role. The researcher and participants had no supervisory or employment relationship, and they did not know each other before recruitment. Contextual observations and researcher assumptions were recorded using detailed field notes and interpretations were regularly checked through team discussions with RH and DN. This minimised individual bias and strengthened analytical rigor.

### 2.6 Data Analysis

We conducted abductive thematic analysis using ATLAS.ti, following the established framework by Braun and Clarke (34,35). This approach allowed us to iteratively move between theory and empirical findings, allowing existing theoretical constructs to guide analysis while remaining open to emerging data patterns.

#### 2.6.1 Coding Approach

An initial deductive codebook was developed from the study’s theoretical lenses. Deductive codes included concepts such as Boundary Discretion, Frugal Improvisation, Information Brokerage, Governance and Coordination Challenges, Intervention Design Challenges and Protocol Friction. This demonstrated how CHPs balance official guidelines with personal judgement when providing maternal care. Inductive coding was conducted concurrently to capture context-specific experiences not fully represented in the theoretical framework. These include Community Hostility and Frustration, Health Access Barriers, Moral Distress, Cultural Frustration, and Dietary Practices that affect their service delivery. Through iterative comparison and iterative refinement, the related codes were grouped into five analytical themes that structure the findings.

**Theme 1: Institutional Fragmentation and System Gaps** captured structural disconnects between health, nutrition, and social protection systems that created practical barriers at household level. This theme brought together codes related to governance challenges, intervention design limitations, protocol friction, health access barriers and failure of formal pathways to connect vulnerable households with appropriate support.

**Theme 2: Workforce Agency and Frontline Adaptation** illustrated how CHPs go beyond their beyond formal roles. By using boundary discretion, creative problem solving, information brokerage, and discretionary practices, they bridge gaps to make households get the care they need.

**Theme 3: Workforce Cost Absorption** captures the financial, operational, relational, and emotional costs CHPs absorb when formal systems inadequately meet reality and service needs. This theme emerged from codes related to out-of-pocket expenditure, personal resource use, additional follow-up time, transport and communication costs.

**Theme 4: Relational and Emotional Burdens** captured the moral distress, frustration and emotional toll that arises from repeated exposure to poverty, trauma, scarcity, community hostility, and situations CHPs could not resolve.

**Theme 5: Cultural, Gendered and Contextual Constraints** captured how socio-cultural norms, gender dynamics, age, hierarchies and contextual factors i.e. urban poverty, pastoral mobility, arid geography shaped household health behaviours and the additional relational work CHPs undertook to negotiate access and adherence.

#### 2.6.2 Advanced Analytical Mapping

Beyond the thematic analysis, three analytical procedures within ATLAS.ti were employed to examine relationships between codes and themes across the dataset. These include co-occurrence tables, code-document comparisons, and network diagrams.

##### 2.6.2.1 Code Co-occurrence Analysis

The co-occurrence matrix (Fig 1) empirically illustrates the relationship between system failures and the personal, physical and financial costs absorbed by the CHPs. The observed patterns also illustrate a process of cost transfer where deficiencies in governance, program design and service delivery were redistributed to the CHPs. The analysis revealed that when top-down infrastructure experienced protocol friction or governance deficits, the vacuum created triggered localised workarounds, with financial subsidisation deeply intertwined with moral distress and boundary discretion.

**Figure. 1.**
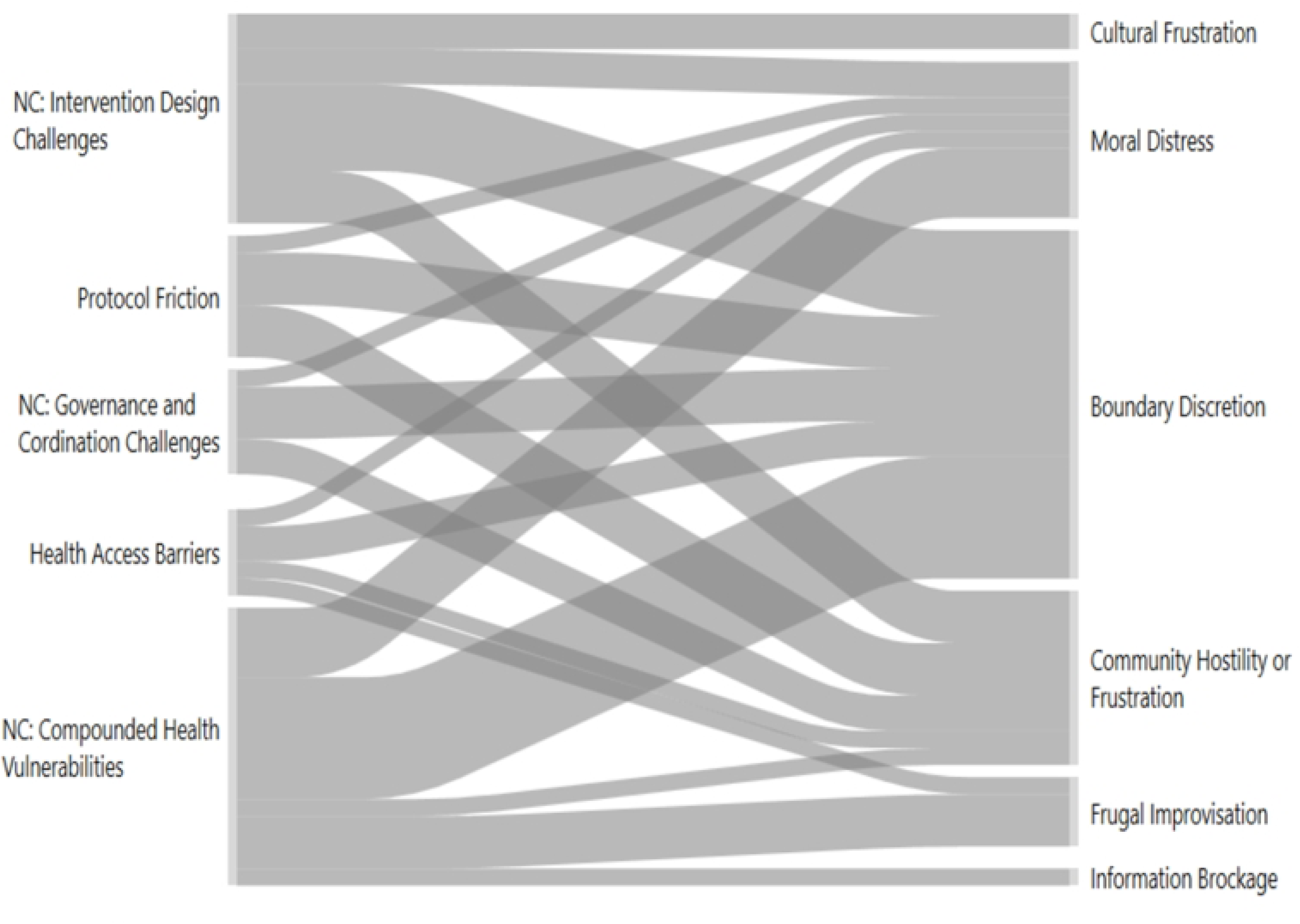

#### 2.6.2 Code-Document Analysis

The code-document thematic distribution (Fig 2) revealed how operational breakdowns manifested differently across the three study areas. Street-level bureaucracy, a key component of Theme 1, emerged as a key theme across the study sites. This indicates that governance challenges, protocol friction, and intervention design challenges were pervasive regardless of the geographic setting. Workforce Agency was similarly evident across counties, reflecting a widespread reliance on adaptive practices. System disconnect distribution differed more noticeably across settings with geographical isolation and logistical barriers associated with nomadic and Arid and Semi-Arid Lands (ASAL) populations. Urban slum challenges were linked to the complexity of densely populated informal settlements.

**Figure. 2.**
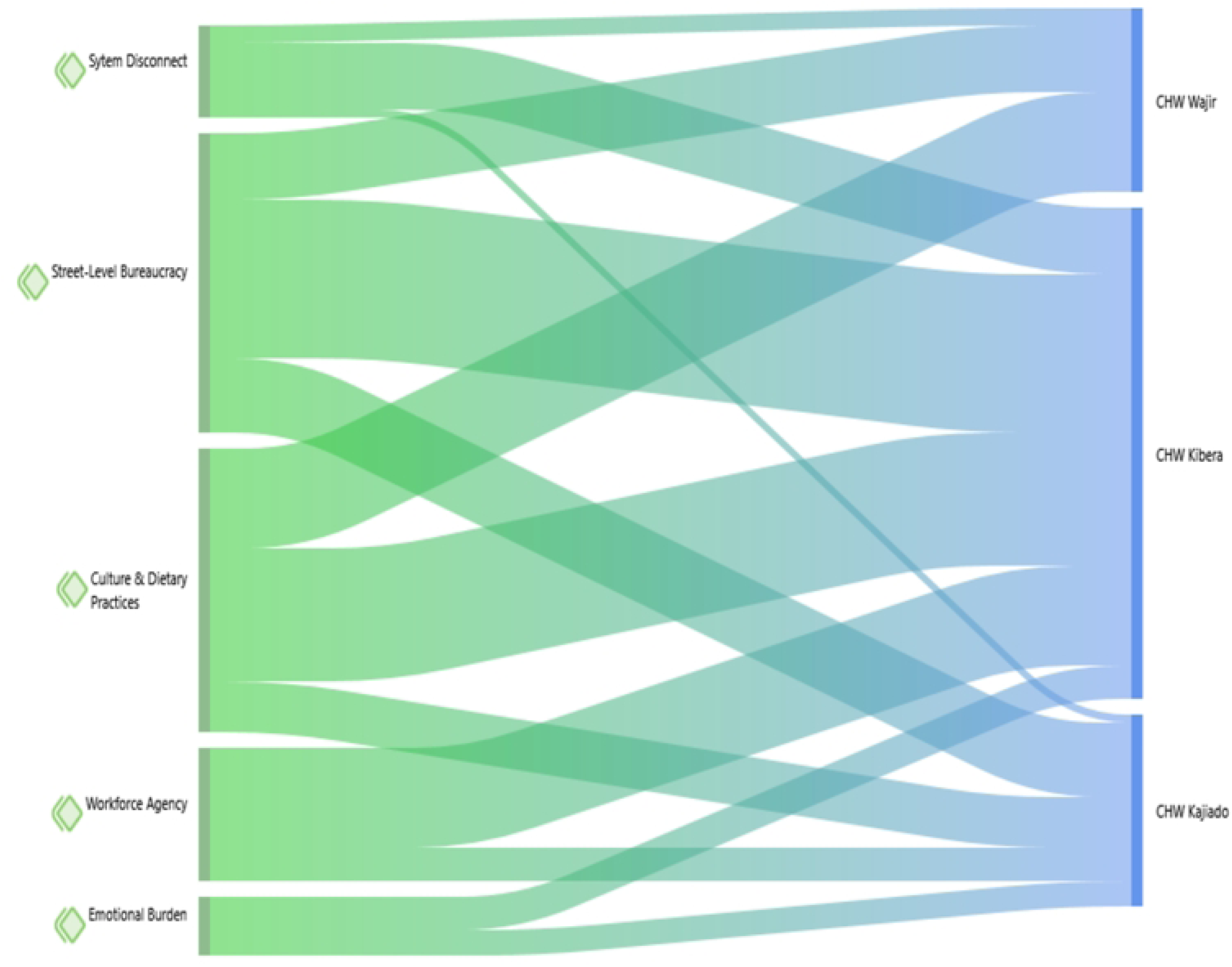

#### 2.6.3 Invisible Safety Net Network

The invisible safety net network (Fig 3 and 4) illustrated the hidden mechanisms keeping maternal nutrition services functioning despite systemic challenges. The informal activities-information brokerage, navigating cultural barriers, exercising boundary discretion and absorbing emotional and ethical burdens were absent in job descriptions, monitoring indicators and funding allocations yet were essential for implementation. The network showed that system failures forced CHPs to do more than their actual jobs activating workforce agency and increasing emotional burden. Additionally, cultural beliefs surrounding pregnancy, dietary practices, and food supplementations influence both maternal health behaviours and CHP decision-making requiring continuous negotiation between nutrition intervention recommendations and community realities, reinforcing the adaptive nature of frontline implementation.

**Figure. 3.**
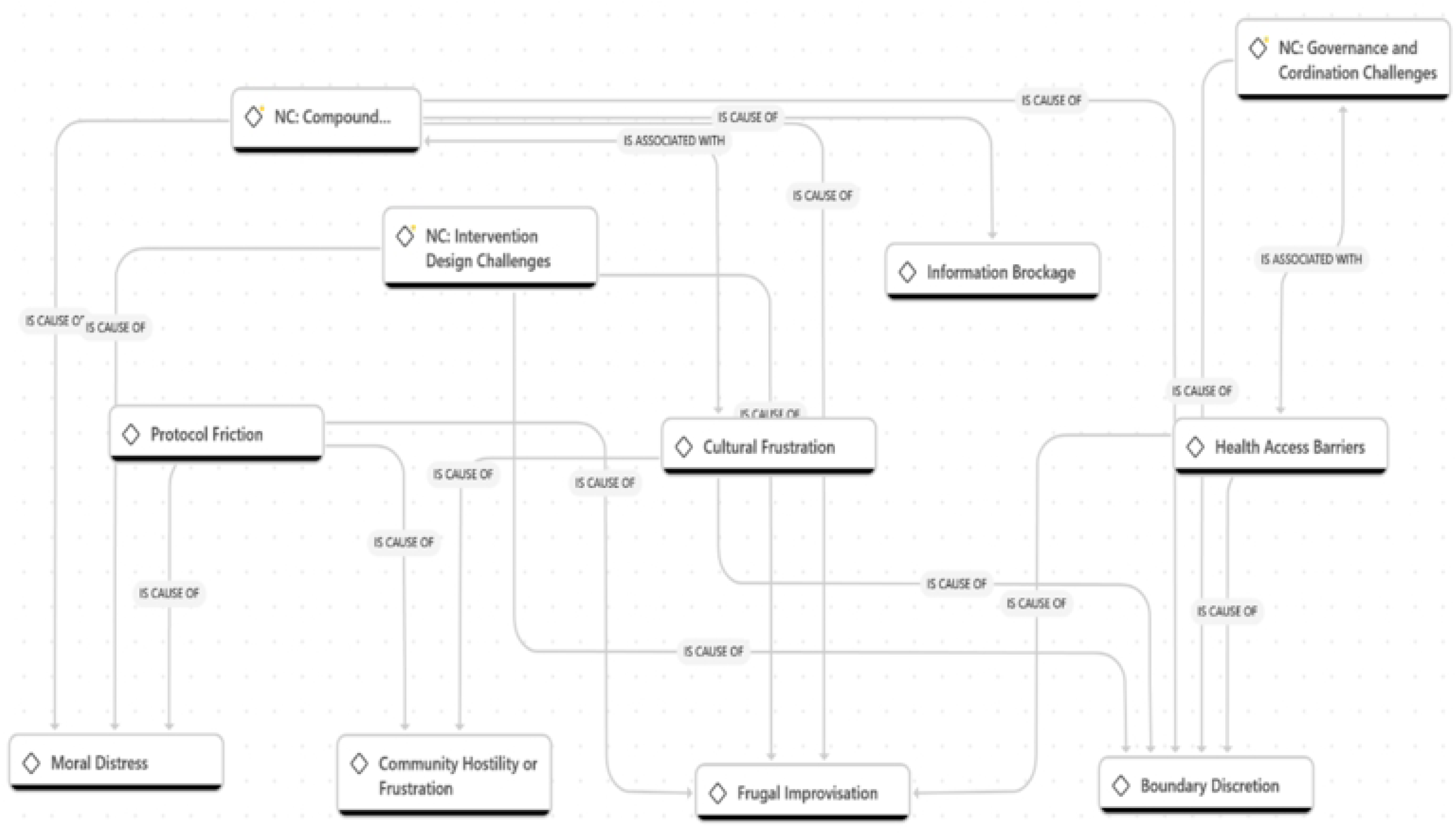

**Figure. 4.**
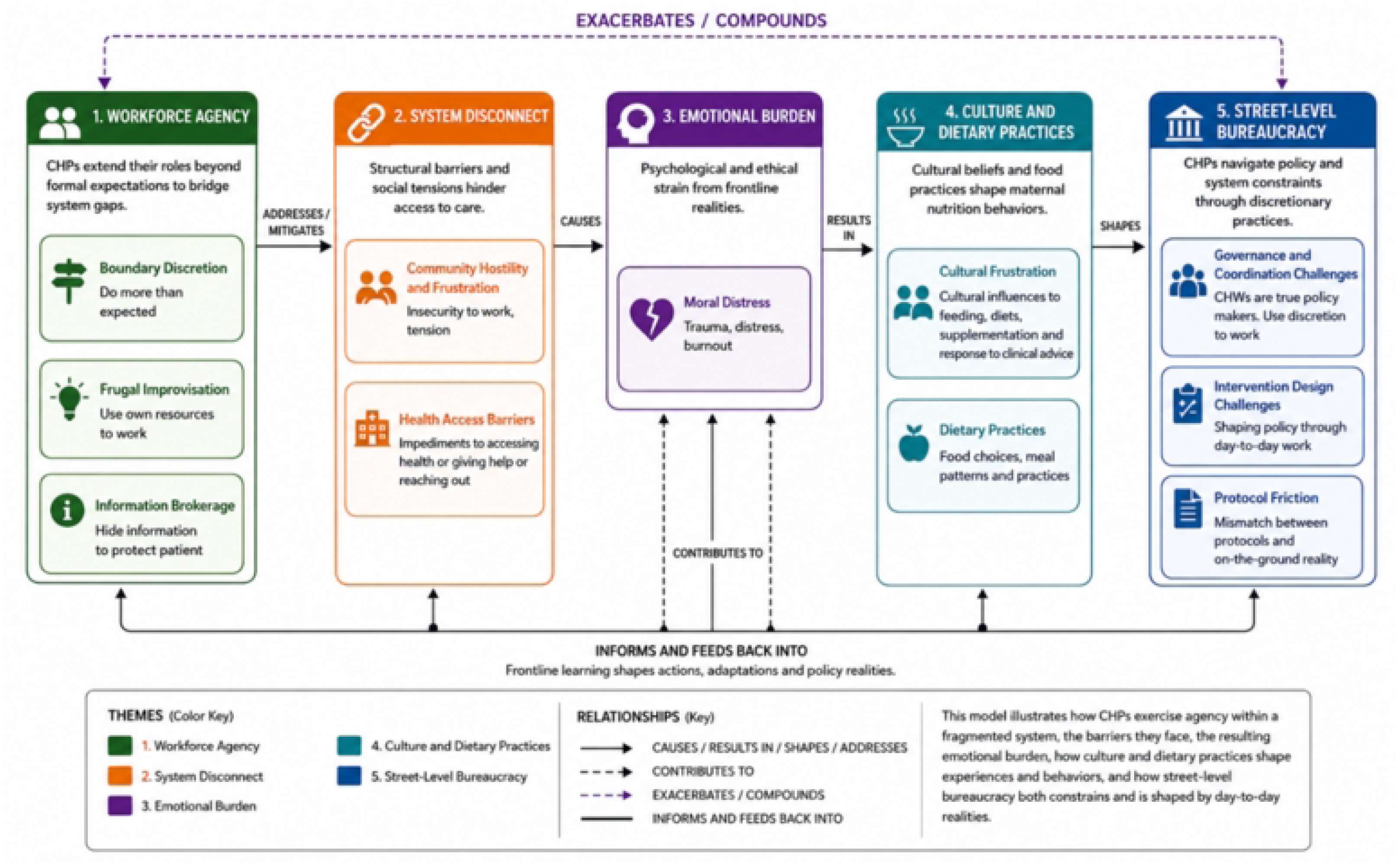

### 2.7 Reporting Standard

The manuscript was prepared following the Consolidated Criteria for Reporting Qualitative Research (COREQ) and the completed COREQ checklist is provided as supplementary material.

### 2.8 Ethics Approval and Consent to Participate

The study was conducted following the Declaration of Helsinki. Ethical clearance was obtained from the University of South Africa IRB (Ref: 2025_SBL_DBL_036_FA_7345) and The Strathmore University Institutional Scientific Ethics Review Committee (SU-ISERC) (Ref: SU-ISERC3058/25). Subsequently, a formal research permit was issued by the National Commission for Science, Technology, and Innovation (NACOSTI) in Kenya, License No: NACOSTI-P-25-4181418. All participants provided informed consent in writing, prior to data collection, with the consents presented in both English and Swahili language for the non-English speaking participants. Participation was voluntary, and data was anonymised to ensure stakeholder confidentiality. Participant recruitment started on 1^st^ November 2025 and ended on 31^st^ January 2026.

## 3. Results

### 3.1 Participant Characteristics

The study analysed detailed narratives of the 12 CHPs representing urban informal, pastoralist and arid contexts. The demographic distribution is detailed below.

Analysis of the CHP interviews reached thematic saturation, with five themes consistently recurring across participants despite differences in geographic context. The themes and the following quotes illustrate the realities of CHPs as they navigated the maternal nutrition intervention responsibilities in the urban informal settlements, and the semi-arid and arid areas under study.

### 3.2 Key Thematic Findings

#### 3.2.1 Institutional Fragmentation and System Gaps

In all the three settings, CHPs described repeated gaps between official health, nutrition and social protection systems which hindered service delivery to household level. These gaps took the form of households lacking the financial resources to act on health recommendations; referral pathways constrained by transport, distance or administrative boundaries, vulnerable households known to community workers but not connected to appropriate social protection mechanisms or digital reporting systems failing due to connectivity or device challenges. As a result, CHPs directly encountered the consequences of fragmentation because they remained responsible for engaging a household after the formal pathway failed to resolve the problem.

One participant described the difficulty in referring a woman to a facility when a household couldn’t meet associated costs:

***“****When we notice health issues and refer to the hospital, sometimes family members take a back seat, and we end up taking on some bills and responsibilities from our work. This scares us and we sometimes just want to not be involved, and you may miss out on this confusion. You get entangled in family matters**”***. -**KIBCHW2**

Similarly, in maternal nutrition, a participant described encounters with women willing to adhere to nutritional advice but lacking resources to implement it:

***“****Talking to a mother struggling to feed is difficult. Some even get aggressive and ask you to give them those things you are asking her to cook-I cannot afford those things you are asking so if you give me, I will cook’. **”*****- KIBCHW1**

CHPs experience technical friction when using state-mandated platforms because of connectivity limitations, malfunctions and a lack of basic facilitation resources like airtime and data. These disrupt data logging and breaks communication continuity.

***“*** *Sometimes we have problems with the system loading and connectivity. We also need to get airtime and data and sometimes the system just hangs. **”*** **KIBCHW3**

**KAJCHW1** reiterates ***“*** *There is code 40327 we CHWs are expected to use to signal/ send SMS different issues in the community and there are 6 unique codes for reporting different issues and we expect a call back for us to give a description of the issue. At first, we used to get a quick callback but these days they may not even respond or delay in picking up the signals. Some households are too hard to reach and have no salaries; we walk long distances to reach them. With ECHIS, we have data sync challenges. **”***

Hardware disparities worsen digital disconnect. CHPs noted that while they are expected to navigate advanced solutions, some of their own basic devices cannot support the required architecture.

***“****Delayed communication between Community Health Workers and health facilities and some mothers lack phones. Some Community health workers have basic phones and cannot access applications. **”*****- WAJCHW3**

Administrative obstacles lock marginalised populations out of healthcare safety nets. With a lack of foundational identification documents, underage women are not able to register for Social Health Insurance, leaving the CHPs unable to secure clinical and social support for them. CHPs are also restricted by administrative jurisdictions that penalise cross-boundary emergency responses.

***“****Some do not have Identity Documents so they are not registered for Social Health Insurance, without this, they cannot get any help. Sometimes we get cases where we help, but I cannot make a referral if the person is not in my jurisdiction. **”*** -**KIBCHW4**

While the findings emphasise system gaps, participants also described situations where formal mechanisms functioned as intended. When digital systems were operational, CHPs could complete data logging and referrals more efficiently. However, these instances were described as inconsistent and depended on factors outside the CHPs’ control:

***“*** *Sometimes we have problems with the system loading and connectivity. **”*****-KIBCHW3**

*“ There is code 40327 we CHWs are expected to use to signal/ send SMS different issues in the community At first, we used to get a quick callback.”*- **KAJCHW1**

*“We use the ECHIS system…it is a good way to keep records.”* -**KIBCHW2**

#### 3.2.2 Workforce Agency and Frontline Adaptation

When formal pathways are insufficient, CHPs adapt their work through discretion, improvisation and informal leadership. They described extending responsibilities through interpersonal relationships, providing additional information and finding alternative ways to maintain communication, more than routine health education. This involved brokerage between households and systems.

One participant described additional effort required when pregnant women concealed their circumstances. Discretion often made incomplete pathways workable:

***“****These mothers sometimes hide their pregnancy, and we have many teen pregnancies in the slum, so I understand when they want to hide the pregnancy and any information around it. We must work extra hard to build trust for them to open up*.

The expectant women have a book where their pregnancy information, progress and everything is recorded, and this is a reference for us, but some hide the books, some lose them intentionally, and some refuse to give the books when asked. Some even tell you it got lost, or I left it at home (the village or up country), and I have been in a situation where, during a home visit with a nurse I let a nurse know that the lady was uncooperative, she had to force the mother to produce the book.**” - KIBCHW1**

***“****Those mothers or even people in general suffering from HIV sometimes never tell their families about their situation and only open up to the Community Health workers so getting information about how they are doing in times of a crisis is hard as some do not know what the issue is and how they can be helped, This forces you to always be available when they need help and this distracts your routine and can inconvenience your work schedule and information involved. **”***

*Teenage pregnancy is quite an issue here, and this comes with a lot of concealed information because of shame, stigma, etc. Some do not want to go back to school, or some get abandoned by their parents, and this makes it hard to follow up on such cases.**”*** **- KIBCHW4**

CHPs described the need to work around community mistrust and suspicion regarding data collection. Some households believed that information collected by CHPs was linked to financial assistance or to CHPs financial gain. CHPs resort to leaving their mobile phones behind, relying on memory to manage suspicion.

***“****Also, going to the houses, you do not carry the phone. When you go with the phone, they have more expectations of you. Also, you do not write a lot, so you have to remember a lot and also be cautious. People think CHWs have money and they feel we owe them.**”*** -**KIBCHW6**

***“****Many assume money is given to CHW, and we use them and the records we collect for our benefit. **”*** -**KIBCHW1**

#### 3.2.3 Workforce Cost Absorption

CHPs described contributing their own resources or absorbing additional operational demands, including paying hospital bills and transport costs out-of-pocket, buying food for starving clients, and using personal airtime and data for follow-up.

***“****When I visit a household, even my colleagues need to carry something for them to even have a place to start my conversation, some food, sugar etc., to encourage them and this is at our own cost.**”*** -**KAJCHW1**

***“****We Community health workers are the ones carrying a huge burden in the community, people reach out to us, and some assume we are meant to be treating them. We understand the issues in the community and have had to sort out some issues at our cost. **”*** **-KIBCHW3**

***“****As CHWs, we also need to visit more, and this means money. **”*** **KAJCHW1**

These contributions illustrate that when formal resources are insufficient, individual workers sometimes absorb part of the resulting cost. Workforce cost absorption was not limited to personal expenditure. Participants also described the consequences of large household allocation, geographical dispersion, and inadequate logistical support. One participant highlighted that CHPs could not always maintain the frequency of contact vulnerable households require.

***“****.I have 100 households and it is not easy to visit all of them. The places are scattered; I do not have transport and sometimes I have to take care of the people myself. Some places are so hard to reach and sometimes you need more help to do your routine checks because we encounter expectant and sickly mothers who sometimes need referral or facilitation to get to the hospital. Also, some people need more time and more frequent visitation, and this makes it difficult to visit all the households as expected so we may miss information due to missed visits or delays. **”*** **KIBCHW3**

***“****Managing 100 households means you visit one every three months and can miss a lot of information especially for pregnant mothers**.”*****- KIBCHW1**

The trade-off between competing needs shows that formal allocations that seem manageable at program level can translate into difficulty making prioritisation decisions at the household level, causing care rationing. CHPs also described digital and administrative friction as a cause of additional costs. Challenges with connectivity, airtime, and devices created extra work.

***“****We use the ECHIS system, but we use our own airtime for a lot of follow-up. **”*** -**KIBCHW2**

In an extreme illustration of the pressure CHPs face, one participant described unauthorised appropriation of hospital supplies, illustrating the moral pressure they may experience when confronted with extreme needs:

***“****One day I had to steal some food from the hospital and take it to a mother who could not make it to the hospital and I was frustrated. I stole some food for her-I just told God I am sorry. **”*** **KAJCHW 1**

This narrative should not be viewed as endorsement or an example of normative practice. Rather, it reflects the profound moral pressure CHPs may experience under extreme household needs and limited options. The expression ‘I just told God I am sorry’ indicated recognition of the ethical tension involved.

#### 3.2.4 Relational and Emotional Burden

Participants described the emotional strain associated with repeatedly encountering poverty, unmet health needs, and situations they could not cater for, producing moral tension. One participant described the frustration of advising women about nutrition when the household lacked food:

*“Most of us have taken up CHP roles beyond pay. It weighs heavily in our hearts when doing the job is hard and we ask for support”*.- **KAJCHW1**

Another described the emotional impact of encountering severe deprivation:

I once visited a mother, I just cried and even came to seek mental help here at the hospital. It is too bad to see. *“-* **KAJCHW2**

CHPs reported encountering defensive friction when delivering standard care and nutrition education. The interactions broke down when families could not afford to follow the recommendations, and some responded aggressively or with withdrew dismissively.

***“*** *Some even get aggressive and ask you to give them those things you are asking her to cook-I cannot afford those things you are asking so if you give me, I will cook’. **”*****- KIBCHW1**

***“****Mothers’ ignorance and Negative responses from mothers are draining. **”*****- KIBCHW3**

Deep-rooted social stigma and widespread communal suspicion around digital data extraction further intensified the burden:

***“****HIV positive mothers assume you want to attack their status; they get aggressive and assume their status is discussed so talking about nutrition is not an easy conversation. **”*** -**KIBCHW1**

Community members who did not receive tangible aid concluded that CHPs profited from their information, sometimes falsifying or withholding information during surveillance:

***“****Some believe that we CHWs get money for our benefit from the information we collect during our visits and some will not be cooperative to share true information or share no information at all. **”***.-**KIBCHW4**

**”**The community also thinks I have been given money to sort their problems and claim I use it for my own benefit. **” WAJCHW1**

Exhaustion is linked to expanding responsibilities and minimal institutional support.

***“W****e CHWs do a lot of work, the facilitation and support is not as much, we meet aggressive people in the community, We deal with some health matters that are beyond our capacity training, or knowledge**”-*** **KIBCHW2**

CHPs also described the consequences of delayed and reduced payments, which created demotivation and financial strain:

***“****Also, we get demotivated sometimes we do not get our stipends. We were promised 50% pay by national government and 50% by the county but we inconsistently get 35% from the county and the national government is at 35%. This does not come in consistently and it is discouraging and even draining to follow up. We even have tasks at health facilities in addition to the home visits and patient care and even with the inconsistent pay, there are many rules to get payment that we do not know how to work. For example, I may be told I will not be paid because I have not met a target, which target when I can spend a whole day handling a crisis in one homestead and need to follow-up the next day. **”*** -**KIBCHW2**

#### 3.2.5 Cultural, Gendered and Contextual Constraints

The work of bridging was also shaped by gender, age, poverty, mobility and socio-cultural expectations. Participants described situations in which women faced constraints in decision-making, disclosure of pregnancy, mobility and access to resources. In some settings, cultural practices and reliance on traditional forms of maternal care affected acceptance of facility-based services or clinical recommendations. Therefore, CHPs had to negotiate and explain health recommendations in locally meaningful ways, building relationships and navigating community authority:

***“****Poverty is a big issue here in the slum and mothers eat the little they have. Some single parents struggle when they get tired and can’t work-they cannot provide and sometimes we are forced to chip in when you get a mother is in a bad situation. Expectant mothers with other children choose to feed their children and may have nothing left for themselves. **”*** -**KIBCHW1**

Deep-rooted cultural practices influence clinical interventions. Female Genital Mutilation generates social stigma and traditional beliefs that undermine women’s health:

*“FGM has so much stigma around it and there are myths that push the agenda at the expense of health. It is believed that women who are not circumcised will have unsuccessful children and those not circumcised are considered kids. **”***- **KAJCHW1**

***“****We still have heavy cultural influence among the Maasai community and those in my household do not go to hospital or care facilities. In fact, they still give birth in the house. So many do not take supplements and do not attend clinics. **”*****- KIBCHW6**

Community myths and misconceptions also interfered with nutrition interventions:

***“****Community myths and misconceptions make them ignore intervention and care advice i.e. they believe it is wrong and a curse to use family planning, as one can never give birth, you will get hormonal imbalances, and it would be a curse from God. **”***- **WAJCHW1**

Age-based social hierarchies mediate community acceptance of health information. Older people were respected as knowledge keepers and trusted cultural mediators, which affects how other CHPs are received.

***“****Sometimes they tend to listen to the elderly CHWs, and that determines if they listen and follow. **”-*** **KIBCHW5** Participants also described superficial compliance, where households agreed to shorten the interaction then dismissed the advice:

***“****Upon information sharing, they respect the health worker and agree to what is advised, just to have a short conversation and do not follow and implement what is advised, you leave them saying you are talking nonsense. **”*****WAJCHW1**

Several distinct gender-related dynamics directly influenced CHP work. First, young women often lacked autonomy to make domestic choices. Even when they accepted nutritional advice, they frequently needed permission to implement. Second, adolescent pregnancy routinely triggered stigma and family rejection. This forced CHPs to dedicate significant time to mediate family conflicts and provide emotional support. Third, entrenched traditions such as Female Genital Mutilation (FGM) and early marriages limited women’s personal freedoms. Ultimately, these barriers force CHPs to balance clinical recommendations, strict community expectations, and women’s lived realities.

## Discussion

### 4.1 Principal Finding

The study examined how CHPs navigate gaps between maternal health, nutrition and social protection systems across three Kenyan settings. The findings reveal a pattern: institutional fragmentation generates household-level gaps, and CHPs respond through frontline adaptation. These responses transfer financial, operational, relational and emotional costs to individual workers, a process conceptualised as workforce cost absorption producing an invisible safety net that sustains service continuity despite system constraints. The workforce cost absorption concept does not imply that CHPs systematically finance the health systems. It does not suggest that CHPs’ adaptations prevent systems collapse. Instead, in the context of fragmented service delivery, the concept describes bounded instances in which specific costs generated by institutional fragmentation are transferred to frontline workers.

The three settings revealed both common patterns and important differences. Gaps in referral pathways, digital systems failures, and personal cost absorption were common to all settings.

However, the nature of fragmentation differed. In Kibera, an urban slum, fragmentation manifested as a coordination challenge among multiple Non-Governmental organisations and government programs creating confusion about eligibility and duplication of effort. In Wajir, fragmentation was primarily geographic, with long distances to facilities, poor infrastructure, and seasonal mobility making referrals challenging. In Kajiado, cultural factors interacted with service fragmentation as traditional birth attendants and community elders mediated access in ways formal programs could not recognise. These differences suggest that interventions addressing fragmentation should be locally tailored while addressing the underlying issue: the absence of integrated pathways between health, nutrition and social protection systems.

### 4.2 Institutional Fragmentation Transfers Cost to Frontline Workers

The finding that fragmented health and social protection systems create practical barriers at household level is consistent with broader health systems literature documenting how institutional silos undermine service integration (11–13). Beyond causing inefficiency, our findings reveal that fragmentation directly transfers operational costs to frontline workers. When formal systems fail, CHPs absorb these shortcomings, transforming system gaps into personal burdens. This aligns with Street-level bureaucracy theory which emphasises how frontline workers exercise discretion while implementing policies under resource constraints (17–19). Our findings suggest discretion serves a dual function: it enables productive adaptation, but it transfers institutional inadequacies onto individual workers. This distinction matters for workforce policy, where one should be promoted while the other signals system failure.

### 4.3 Workforce Cost Absorption

The concept of workforce cost absorption contributes to community health worker literature in two ways. First, it distinguishes between workload (volume of assigned tasks) and cost absorption (additional resources required when systems fail). This matters because workload measures alone do not capture the hidden costs borne by CHPs (20–24). Furthermore, it explains a contradiction: the CHP resilience can conceal system flaws. Services appear functional when workers compensate for gaps, reducing pressure for institutional reforms. The feedback loop appear as follows: system gap leads to frontline adaptation, enabling apparent continuity which reduces the gap’s visibility and sustains resilience through adaptation. This extends Schaaf et. Al’s (36) description of CHP as a service extender showing that the extension is not resource-neutral. Taking up the burden, the concept should not be interpreted as suggesting that CHPs systematically finance health systems, rather; it describes a bounded process; in which, some cost fragmentation is transferred to individual workers.

### 4.4 Structural Relational and Emotional Costs

The described emotional burden stemming from community hostility, frustration, and moral distress is not a primary psychological issue but a structural consequence of institutional fragmentation. CHPs face moral tension because they are positioned as the visible face of systems that cannot meet household needs. These findings challenge framings that attribute CHP burnout to resilience deficits. Instead, they suggest that the emotional burden stems from the gap between professional responsibility and institutional capacity. This has implications for workforce support, as psychosocial support for CHPs is necessary but insufficient if the causes are not addressed. As other studies have documented (25,26), failing to address system-level failures individualises systemic problems.

### 4.5 The Need for Structural Responses to Cultural and Gendered Constraints

Literature in sub-Saharan contexts documents that cultural practices, gender dynamics and household power structures shape maternal nutrition engagement (37–39). Our findings not only confirm that these constraints interfere with women’s health but also generate additional work for CHPs, who must deal with structural inequalities; negotiate access, build trust, and mediate between households, power dynamics, and institutions to be effective. This work is invisible in conventional workforce metrics. Gendered powered dynamics leave them to develop individual workarounds that carry personal costs. A household visit may be recorded as one contact, which does not capture the time required to build trust, negotiate disclosures, navigate family dynamics, and address cultural barriers. This invisibility has its consequences: if programs count contacts rather than relational effort, they risk underestimating the work required to engage vulnerable households.

## 4. Implications for Policy and Practice

### 5.1 Overarching Principles

The study findings suggest four policy action points.

First, workforce planning must account for cost absorption. Routine measures of CHP workload should be supplemented with indicators of additional costs incurred when systems fail. Secondly, referral pathways require realistic support mechanisms. A referral is incomplete if the household cannot access the intended destination and referral programs must consider transport, affordability, social protection eligibility and follow-up alongside the referral itself. Third, digital systems should be evaluated by their effect on frontline work. Beyond technical functionality, assessments should examine how the system affects or redistributes work among frontline workers. Fourth, CHPs should not be required to bridge institutional gaps; instead, institutional systems integration must be built around lived realities and household needs, with clear referral pathways, defined responsibilities, accessible eligibility mechanisms, feedback loops, and escalation mechanisms. Finally, while involving CHPs in social protection, safeguard and confidentiality protocols need to be established with accountability mechanisms and workload adjustments to protect both CHPs and communities.

### 5.2 Recommendations

Building on the principles, we propose the following specific actions for different actors within the health and social protection domains.

#### National Government

The national government should formally recognise CHPs as trusted actors in social protection program implementation with structured roles. It should also ensure the 50/50 national and county co-financing agreements are fulfilled consistently. Finally, it should establish a cross-sectoral mechanism such as interoperable information systems, and shared eligibility criteria, to strengthen collaboration between health and social welfare systems, recognising grassroots leadership as a strategic component of the system’s resilience.

#### County Government

County governments should guarantee CHP remuneration by implementing reliable payment systems that do not penalise CHPs for crisis management and performance targets should account for the unpredictable nature of community work. They should also provide sufficient logistical support such as transport costs, data and airtime, and functioning devices. Finally, there is a need for the implementation of psychosocial support such as debriefing, and access to counselling to address the distress arising from their work exposure.

#### Health Facilities

Ensure referral pathways include transport and are affordable. Facility-level protocols need to include support and follow-up mechanisms with linked to social protection. Establish clear escalation mechanisms for CHPs to activate when they identify vulnerable households needing help beyond clinical care.

#### Social Protection Agencies

Involve CHPs in identifying, verifying, and updating beneficiary information, leveraging their proximity to households to reduce exclusion errors. With this, address workload concerns, with the new social protection responsibilities accompanied by appropriate compensation.

#### Program Implementors and Technology Developers

Embrace co-design of cross-sectoral, digital decision support systems around existing CHP workflows, rather than imposing top-down designs, with tools the support informed professional discretion rather than rigid protocol enforcement. Additionally, digital systems should be evaluated by their effect on frontline work and not just technical functionality. The assessment should examine whether the tools reduce or increase CHP workload, whether they are usable with available infrastructure and whether they can strengthen community trust.

## 5. Strengths and Limitations

### Study Strengths

The study’s multi-site approach enabled examination across three contrasting contexts. Focusing on CHPs allowed detailed analysis of the interface between institutional arrangements and household realities. Using advanced mapping techniques such as code co-occurrence and code-documents tabulation allowed rigorous, transparent translation of raw data into complex structural insights.

### Study Limitations

The following limitations should be considered. First, the analysis is based on 12 CHPs from a broader 75 stakeholders; findings are not statistically representative. As a result, interpret the findings as contextually transferable rather than statistically generalizable to all CHPs or other sub-Saharan settings. Secondly, while thematic saturation was achieved, the sample size (n=12) and uneven distribution across sites limit the ability to determine how common the reported practices are across different settings. Third, the findings rely on self-reported accounts and recall, and social-desirability bias is possible. Fourth, the interviews were conducted in both English and Swahili, and translation may have reduced nuance. The first author’s bilingual proficiency mitigated the risk, but some contextual meaning may have been lost. Finally, the claims were not triangulated with maternal accounts and administrative records and the cross-sectional design limits conclusions about changes over time in CHP workload or cost absorption.

## Conclusion

This study shows that CHPs do not just deliver maternal nutrition interventions. They act as vital moral agents and unmapped safety nets for resource-constrained systems. When institutional separation excludes vulnerable mothers, CHPs use personal discretion and absorb costs to bridge the gap. However, relying on this unpaid moral labour is unsustainable, as defunded agreements, heavy quotas, community hostility and constrained digital tools erode worker motivation and community trust.

## Data Availability

The datasets generated and/or analysed during the current study are not publicly available due to the sensitive nature of the interviews, but are available from the corresponding author on reasonable request

## Acknowledgment

We acknowledge The University of South Africa, Strathmore University and NACOSTI for this opportunity. We deeply appreciate the anonymous study participants for their support and invaluable insights, and for sharing their service realities.

## Supporting Information

## Declarations

## Consent for Publication

Not Applicable

## Authors Contribution

FS conceptualised the study, collected data, conducted analysis and drafted the manuscript. RH and DN provided methodological supervision, critically reviewed the analysis and contributed to the interpretation of the findings,. They approved the final manuscript.

